# Low and Borderline Ankle Brachial Index is Associated with Intracranial Aneurysms – A Retrospective Cohort Study

**DOI:** 10.1101/2023.07.11.23292533

**Authors:** Dan Laukka, Essi Kangas, Aino Kuusela, Jussi Hirvonen, Tiia Rissanen, Melissa Rahi, Juri Kivelev, Ville Rantasalo, Jaakko Rinne, Harri Hakovirta

## Abstract

**Background and Aims:** A low ankle-brachial index (ABI) is associated with a higher risk of cardiovascular diseases, stroke, and systemic inflammation. Intracranial aneurysms (IAs) share similar risk factors with other cardiovascular diseases. However, the association between low ABI and IAs lacks sufficient investigation.

**Methods:** This retrospective study reviewed 2751 patients who had ABI measurements at a tertiary hospital from January 2011 to December 2013. Patients with available cerebrovascular imaging or a diagnosis of ruptured IA were included in the study (n=776) to examine the association between ABI and saccular IAs. The patients were classified into four groups: low ABI (≤0.9, n=464), borderline ABI (0.91-0.99; n=47), high ABI (>1.4, n=57), and normal ABI (1.00-1.40; n=208).

**Results:** The prevalence of IAs was 20.3% (18.1% unruptured IAs) in the low ABI group, 14.9% (12.8% unruptured IAs) in the borderline ABI group, 7.0 % (5.3% unruptured IAs) in the high ABI group, and 2.4% (1.9% unruptured IAs) in the normal ABI group (*p<*0.001). Sex- and age-adjusted multinomial regression, including clinically relevant variables, revealed that low ABI (odds ratio [OR], 11.33; 95% confidence interval [CI], 4.08-31.51; *p<*0.001) and borderline ABI (0.91-0.99) (OR, 7.13; 95% CI, 1.91-26.63; *p=*0.004) were the only variables significantly associated with unruptured IAs.

**Conclusions:** The prevalence of unruptured IAs was 9-fold higher in the low ABI group and nearly 7-fold higher in the borderline ABI group when compared to the normal ABI group. ABI measurements could be clinically relevant for identifying individuals at higher risk of IAs and may help guide screening and preventive strategies.

## 1. Introduction

The prevalence of unruptured intracranial aneurysms (IAs) in the general population is approximately 3%,^1^ and the annual incidence of aneurysmal subarachnoid hemorrhage is approximately 8 per 100 000^2^. Hypertension and smoking are the most important modifiable risk factors for IAs.^3, 4^ Higher atherosclerotic burden may be associated with an increased risk of IAs ^5, 6^, and ruptured IAs may be linked to excess mortality due to cardiovascular diseases^7, 8^.

The ankle-brachial index (ABI) is an easily available method for confirming the diagnosis of peripheral artery disease. An ABI ≤0.9 and >1.4 is considered abnormal, whereas ABIs falling within a range of 0.91-0.99 are considered borderline.^9^ A low ABI (≤0.9) and a borderline ABI are associated with an increased risk of cardiovascular diseases and stroke.^10, 11^ ABI can also be used to assess cardiovascular disease risk independently of other risk factors (Stone et al., 2022).^12^ A low ABI may associated with an increase in the risk of abdominal aortic aneurysms,^13, 14^ whereas aortic aneurysms may increase the risk of cerebral aneurysms^15^ and vice versa^16^.

Despite the known predictive value of ABI for cardiovascular disease and potentially for aortic aneurysms, there have been no published studies that have investigated the association between ABI and IAs as far as the authors are aware. In this study, we present an association between ABI and IAs.

## 2. Methods

The study was approved by the institutional review board at Turku University Hospital, Finland. Patient consent was not required because of the retrospective nature of the study.

### 2.1. Population

All consecutive patients (n=2757) who had ABI determined in the vascular laboratory at the Department of Clinical Physiology, Turku University Hospital, South-West Finland, from 1 January 2011 to 31 December 2013 were reviewed retrospectively. The vascular laboratory provides non-invasive pressure measurement that cover the hospital region catchment area population of 480 000 inhabitants.

Radiological examinations and electronic patient charts were reviewed until 1 January 2023. Of the 2757 patients, 776 with available imaging studies (MR angiography, CT angiography or digital subtraction angiography) or a history of ruptured IA were included.

### 2.2. Baseline measurements

The following variables were obtained from the electronic patient records: hypertension (if the patient had diagnosed hypertension and/or anti-hypertensive medication); hypercholesterolemia (if the patient had diagnosed hypercholesterolemia and/or medication for hypercholesterolemia); diabetes type 1; diabetes type 2 (diagnosed diabetes type 2 and/or medication for diabetes type 2); coronary artery disease (if the patient had diagnosed coronary artery disease, prior artery bypass surgery or angioplasty, or prior myocardial infarction); malignancy (prior diagnosis of any malignancy); chronic obstructive pulmonary disease (if the patient had diagnosed chronic obstructive pulmonary disease); malignancy (if the patient had a history of any malignancy); rheumatoid arthritis (if the patient had diagnosed rheumatoid arthritis); varicose ulcus (if the patient had diagnosed varicose ulcus).

Qualified sonographers measured ankle and brachial systolic blood pressure ipsilaterally for both sides of the patient using the oscillometric method. The ABI was calculated as the ratio of the ankle systolic blood pressure divided by the highest systolic blood pressure of the arm ipsilaterally. The lower value of the bilateral ABI determinations was used for analysis.

One of the authors (D.L., Consultant Neurosurgeon) evaluated all cerebrovascular imaging studies for intracranial aneurysms (IAs). A neuroradiologist, with over 10 years of experience, evaluated every IA to confirm the diagnosis. Any disagreements in the evaluations between the neurosurgeon and neuroradiologist were resolved through consensus.

IAs that were saccular and larger than or equal to 2 mm in size and located intracranially were considered as an IA.

According to the Bouthillier classification^17^, aneurysms located distal to the clinoid segment (C5) were defined as intradural, whereas aneurysms located in the intracavernous segment (C4) were classified as extradural. Aneurysms in and proximal to the C4 segment were not analyzed. Unruptured IAs located in the internal carotid artery (ICA) were categorized as follows: ophthalmic artery, posterior communicating artery, anterior choroidal artery, and ICA bifurcation aneurysms. Middle cerebral artery aneurysms (MCA) were categorized as M1-segment, M1 bifurcation, M1/M2 bifurcation, M2, and M3-M4 segments. Anterior cerebral artery aneurysms (ACA) were categorized as A1-segment, anterior communicating artery, and distal ACA aneurysms. Finally, posterior circulation artery aneurysms (PCA) were categorized as posterior cerebral artery P1, P2, and P3, basilar tip, basilar trunk, superior cerebellar artery, posterior inferior cerebellar artery, and anterior inferior cerebellar artery.

### 2.4. Statistical methods

The primary outcome examined was the prevalence of both unruptured and ruptured IAs, either together or separately.

Patients were classified into four main groups based on their ABI values as follows: low ABI (≤0.9), borderline ABI (0.91-0.99), normal ABI (1.0-1.4), and high ABI (>1.4). Furthermore, patients were categorized into three groups, namely: those with ruptured IAs, those with unruptured IAs, and those without IAs.

Associations between the different groups were calculated using the chi-square test for categorical data. For continuous data, Kruskall-Wallis test were used for nonparametric data and one way ANOVA for normally distributed data.

Multinomial regression analyses were performed to evaluate the association of ABI groups with the IAs. In the first set of calculations, adjustments were made for clinically relevant variables. In the second set of calculations, only variables that showed statistically significant values (*p<*0.05) in the univariate analysis were included in the multivariate analysis. Similarly, the associations between ABI groups and covariates were calculated. Both models were adjusted for sex and age.

Continuous data are expressed as either the mean ± standard deviation (SD) or the median interquartile range (IQR) depending on their distribution. Missing data were excluded from the analysis.

Data were analyzed using the JMP16 statistics package (SAS Institute, Cary, NC, USA). A p-value below 0.05 was considered statistically significant.

## 3. Results

Of the 776 included individuals 464 (59.8%) had a low ABI (≤0.9), 47 (6.1%) had a borderline ABI (0.91-0.99), 57 (7.3%) had a high ABI (>1.4), and 208 (26.8%) had a normal ABI (1.0-1.4). Regarding cerebrovascular imaging analyses, there were no disagreements regarding the diagnosis of IAs between the two interpreters.

### 3.1. Associations of ABI groups with intracranial aneurysms

**Table 1** presents the main baseline demographics of patients and intracranial aneurysms (IAs) categorized by ABI groups. **Figure 1** presents the prevalence of unruptured and ruptured IAs by ABI groups.

**Table 1.** Comparison of baseline and intracranial aneurysm (IA) characteristics according to the ABI group. Bold indicate statistically significant value *p<*0.05.

| **Baseline characteristics** | **Normal ABI**<br>**(1.0-1.4)**<br>***n* = 208** | **Low ABI**<br>**($\leq 0.9$)**<br>***n* = 464** | **Borderline ABI**<br>**(0.91-0.99)**<br>***n* = 47** | **High ABI**<br>**($> 1.4$)**<br>***n* = 57** | **p-value** |
| --- | --- | --- | --- | --- | --- |
| Mean age at ABI measurement, years, SD | 67.0 ± 10.8 | 69.8 ± 9.6 | 66.1 ± 13.1 | 68.3 ± 11.5 | **0.003** |
| Mean age at cerebrovascular imaging, years, SD | 69.4 ± 11.9 | 72.2 ± 10.0 | 68.6 ± 14.2 | 68.0 ± 12.6 | **0.010** |
| Sex (female) (%) | 39.4% | 38.4% | 53.2% | 22.8% | **0.016** |
| Comorbidities |  |  |  |  |  |
| Hypertension (%) | 60.1% | 66.0% | 59.6% | 75.4% | 0.129 |
| Diabetes type 1 or 2 (%) | 35.1% | 36.9% | 46.8% | 63.2% | **<0.001** |
| *Diabetes type 1 (%)* | 7.2% | 3.9% | 19.2% | 15.8% | **<0.001** |
| *Diabetes type 2 (%)* | 27.9% | 33.3% | 25.5% | 50.9% | **0.008** |
| Hypercholesterolemia (%) | 24.0% | 30.5% | 27.7% | 36.8% | 0.19 |
| Coronary artery disease (%) | 29.8% | 36.0% | 34.0% | 42.1% | 0.27 |
| Chronic heart failure (%) | 13.9% | 16.4% | 21.3% | 28.1% | 0.069 |
| Atrial fibrillation | 26.0% | 19.4% | 29.8% | 36.8% | **0.008** |
| Chronic kidney failure (%) | 10.6% | 11.9% | 19.2% | 33.3% | **<0.001** |
| Chronic obstructive pulmonary disease (%) | 6.3% | 14.7% | 12.8% | 14.0% | **0.022** |
| Rheumatoid disease | 11.1% | 7.3% | 6.4% | 10.5% | 0.371 |
| Varicose_ulcus | 7.2% | 5.6% | 8.5% | 14.0% | 0.112 |
| Malignacy | 17.8% | 19.4% | 12.8% | 31.6% | 0.070 |
| Intracranial aneurysm characteristics |  |  |  |  |  |
| Prevalence of intracranial aneurysm, total, *n* (%) | 5 (2.4%) | 94 (20.3%) | 7 (14.9%) | 4 (7.0%) | **<0.001** |
| *Unruptured, n (%)* | 4 (1.9%) | 84 (18.1%) | 6 (12.8%) | 3 (5.3%) | **<0.001** |
| *Ruptured, n (%)* | 1 (0.5%) | 10 (2.2%) | 1 (2.1%) | 1 (1.7%) | 0.277 |
| Median size of the largest IA, mm (IQR) | 4.0 (3.0-5.0) | 4.0 (3.0-5.0) | 3.0 (3.0-8.0) | 4.0 (3.0-7.0) | 0.929 |
| Distribution of largest IA |  |  |  |  | 0.817 |
| *Anterior cerebral artery* | 20.0% | 25.8% | 28.6% | 50.0% |  |
| *Internal Carotid Artery* | 60.0% | 31.2% | 14.3% | 25.0% |  |
| *Middle Cerebral Artery* | 20.0% | 21.5% | 28.6% | 0% |  |
| *Posterior Circulation* | 0% | 8.6% | 28.6% | 0% |  |
| *Intracavernous Internal Carotid Artery* | 0% | 12.9% | 0% | 25.0% |  |

**Figure 1.**
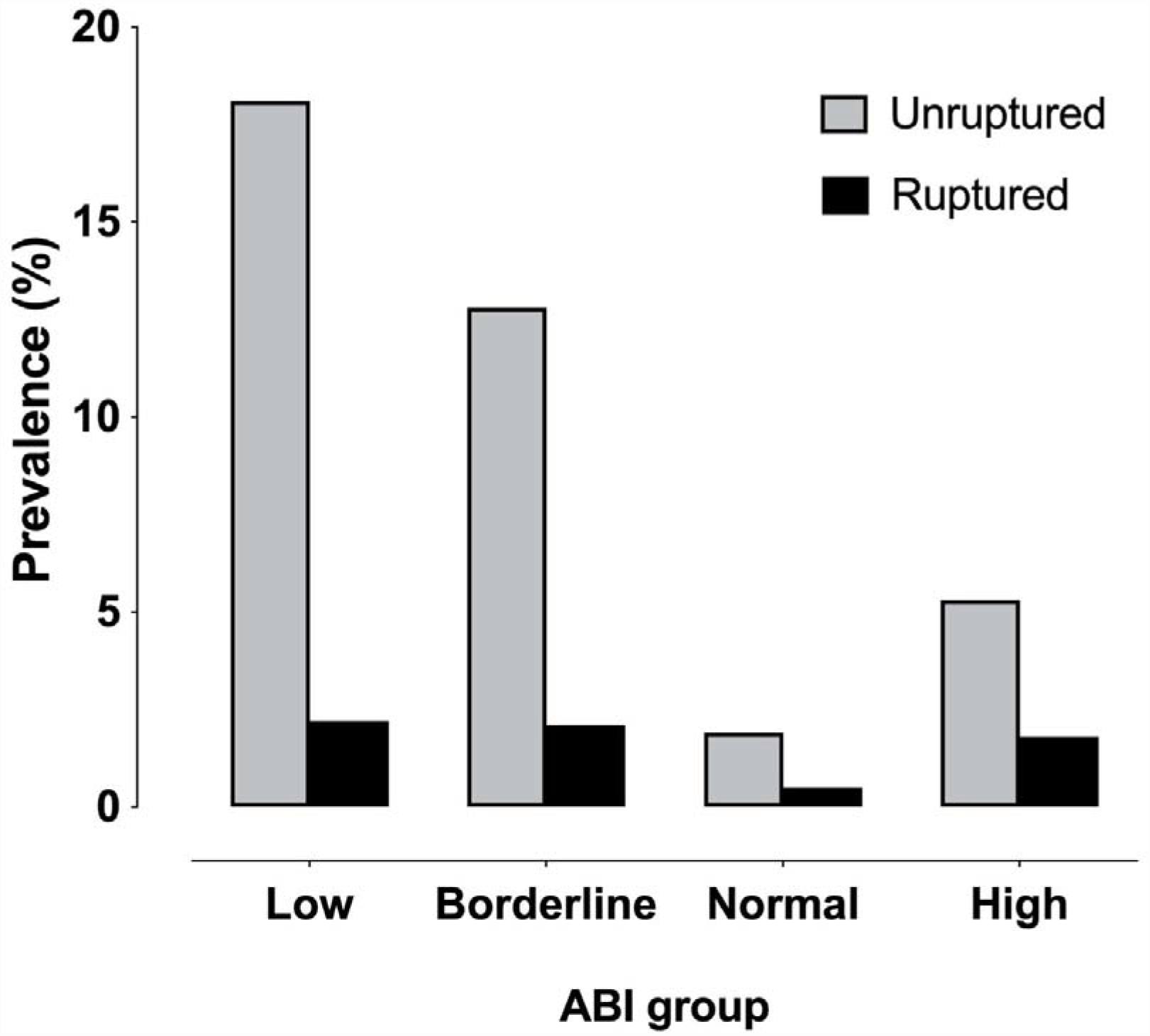
Prevalence of unruptured and ruptured intracranial aneurysms by ankle-brachial index group.

The prevalence of unruptured IAs was 18.1% in the low ABI (≤0.9) group, 12.8% in the borderline ABI (0.91-0.99) group, 5.3% in the high ABI (>1.4) group, and 1.9% in the normal ABI (1.0-1.4) group (*p<*0.001).

Within the low ABI (≤0.9) group, 2.2% had ruptured IAs, whereas in the borderline ABI (0.91-0.99) group, 2.1% had ruptured IAs. In the high ABI (>1.4) group, 1.8% had ruptured IAs, and in the normal ABI (1.0-1.4) group, 0.5% had ruptured IAs (*p=*0.277).

**Table 2** displays the main baseline demographics of patients categorized by IA presentation. The median ABI was 0.59 (IQR 0.45-0.75) in patients with unruptured IAs, 0.57 (IQR 0.51-0.79) in patients with ruptured IAs, and 0.80 (IQR 0.53-1.13) in patients without unruptured IAs (*p<*0.001).

**Table 2.** Baseline characteristics of unruptured and ruptured intracranial aneurysms (IA) and patients without IAs. Bold indicate statistically significant value *p<*0.05.

|  | **Unruptured IA**<br>**n= 97** | **Ruptured IA**<br>**n= 13** | **Without IA**<br>**n= 666** | **p-value** |
| --- | --- | --- | --- | --- |
| Age, years, mean (SD) | 69.5 (8.7) | 68.9 (8.9) | 68.6 (10.6) | 0.753 |
| Sex (male) (%) | 56.7% | 53.9% | 62.5% | 0.467 |
| Hypertension (%) | 68.0% | 76.9% | 64.0% | 0.477 |
| Diabetes type 1 or 2 (%) | 39.2% | 15.4% | 39.4% | 0.213 |
| *Diabetes type 1 (%)* | 5.2% | 0% | 6.9% | 0.508 |
| *Diabetes type 2 (%)* | 34.0% | 15.4% | 32.8% | 0.396 |
| Hypercholesterolemia (%) | 34.0% | 30.8% | 28.3% | 0.508 |
| Coronary artery disease (%) | 29.9% | 7.7% | 35.9% | 0.061 |
| Chronic heart failure (%) | 19.6% | 0.0% | 16.8% | 0.207 |
| Atrial fibrillation | 16.5% | 7.7% | 24.3% | 0.096 |
| Chronic kidney failure (%) | 15.5% | 15.4% | 13.2% | 0.820 |
| Chronic obstructive pulmonary disease | 18.6% | 7.7% | 11.4% | 0.118 |
| Rheumatoid disease | 6.2% | 23.1% | 8.6% | 0.121 |
| Varicose ulcer | 6.2% | 23.1% | 6.6% | 0.064 |
| Malignancy | 17.5% | 7.7% | 20.0% | 0.475 |
| ABI, median (IQR) | 0.59 (0.45-0.75) | 0.57 (0.51-0.79) | 0.80 (0.53-1.13) | **<0.001** |

Multinomial regression analyses, adjusted for age, sex, and clinically significant variables (**Table 3, Model 1)**, revealed that low ABI (≤0.9) (odds ratio [OR], 11.33; 95% confidence interval [CI], 4.08-31.51; *p<*0.001) and borderline ABI (0.91-0.99) (OR, 7.13; 95% CI, 1.91-26.63; *p=*0.004) were the only variables associated with unruptured IAs. Similar results were observed in the second model (**Table 3, Model 2)**, which were adjusted for age, sex and statistically significant variables.

**Table 3.** Multinomial regression models for unruptured intracranial aneurysms adjusted with sex- and age. In Model 1 clinically significant variables and in Model 2 variables that were statistically significant in the univariate analysis. Bold indicate statistically significant value *p<*0.05.

| Variable | Odds ratio | 95% CI | *p* Value |
| --- | --- | --- | --- |
| <u>Model 1</u> |  |  |  |
| Age | 1.00 | 0.98-1.02 | 0.946 |
| Sex (female vs. male) | 1.27 | 0.80-1.99 | 0.309 |
| Hypertension | 1.17 | 0.72-1.90 | 0.516 |
| Coronary artery disease | 0.68 | 0.42-1.11 | 0.120 |
| Chronic obstructive pulmonary disease | 1.52 | 0.84-2.72 | 0.163 |
| Chronic kidney failure | 1.33 | 0.71-2.52 | 0.376 |
| Normal ABI 0.9-1.4 | ref. |  |  |
| Low ABI $\leq 0.9$ | 11.33 | 4.08-31.51 | **<0.001** |
| Borderline ABI | 7.13 | 1.91-26.63 | **0.004** |
| High ABI $> 1.4$ | 2.73 | 0.58-12.74 | 0.473 |
| <u>Model 2</u> |  |  |  |
| Age | 0.99 | 0.98-1.02 | 0.951 |
| Sex (female vs. male) | 1.28 | 0.82-1.99 | 0.286 |
| Normal ABI 0.9-1.4 | ref. |  |  |
| Low ABI $\leq 0.9$ | 11.60 | 4.18-32.17 | **<0.001** |
| Borderline ABI | 7.38 | 1.99-27.95 | **0.003** |
| High ABI $> 1.4$ | 3.00 | 0.65-13.84 | 0.160 |

### 3.2. Associations with ABI groups

The multinomial regression analysis revealed age (OR, 1.03; 95% CI, 1.02-1.05; *p<*0.001), chronic obstructive pulmonary disease (OR, 2.68; 95% CI, 1.43-5.01; *p=*0.002), and atrial fibrillation (OR, 0.53; 95% CI, 0.35-0.80; *p*<0.001) were associated with low ABI (≤0.9). Diabetes type 1 or 2 (OR, 2.41; 95% CI, 1.26-4.61; *p=*0.010) and chronic kidney failure (OR, 3.29; 95% CI, 1.55-6.96; *p=*0.002) were associated with high ABI (>1.4). Female sex (OR, 1.95; 95% CI, 1.02-3.73; *p=*0.04) was associated with borderline ABI (0.91-0.99). Refer to **Table 4** for further details.

**Table 4.** Risk factors for Low ABI (≤0.9), Borderline ABI (0.91-0.99), and High ABI (>1.4) compared to normal ABI (1.0-1.4). Model included variables that were statistically significant in univariate analysis. Values are odds ratios (95% CI). Bold indicate statistically significant value *p<*0.05.

| Variable | Low ABI | Borderline ABI | High ABI |
| --- | --- | --- | --- |
| Age | **1.03 (1.02-1.05)** | 0.99 (0.96-1.02) | 1.02 (0.99-1.05) |
| Sex (female vs. male) | 0.99 (0.70-1.40) | **1.95 (1.02-3.73)** | 0.53 (0.26-1.06) |
| Diabetes type 1 or 2 | 1.17 (0.81-1.68) | 1.58 (0.81-1.68) | **2.41 (1.26-4.61)** |
| Atrial fibrillation | **0.53 (0.35-0.80)** | 1.26 (0.60-2.66) | 1.46 (0.75-2.86) |
| Chronic kidney failure | 1.18 (0.68-2.05) | 1.58 (0.81-3.10) | **3.29 (1.55-6.96)** |
| Chronic obstructive pulmonary disease | **2.68 (1.43-5.01)** | 2.45 (0.87-6.92) | 2.86 (0.87-5.98) |

## 4. Discussion

We found a significant association between low ABI (≤0.9) and between borderline ABI (0.91-0.99) with an increased risk of IAs when compared to patients with a normal ABI (1.0-1.4). The prevalence of unruptured IAs in the low ABI group (≤0.9) was 18.1%, whereas the prevalence of ruptured IAs was 2.2%. Similarly, in the borderline ABI group, the prevalence of unruptured IAs was 12.8%, and ruptured IAs were found in 2.1% of the cases. In contrast, individuals with a normal ABI had a much lower prevalence of unruptured IAs (1.9%) and ruptured IAs (0.5%). This pattern of findings suggests a vastly increased risk of IAs in patients with reduced ABI.

Our findings demonstrate a 9-fold higher prevalence of unruptured IAs in the low ABI group and approximately 6-fold higher prevalence in the borderline ABI group compared to those with a normal ABI range of 1.0-1.4. Notably, the prevalence of unruptured IAs in the normal ABI group was similar to that reported in the general population.^1^

The prevalence of unruptured IAs in the low ABI group and borderline ABI group was similar to that observed in specific populations known to have a high risk of IAs, such as patients with polycystic kidney disease or those who have at least two first-degree relatives with IAs^18^. Consequently, screening for IAs is recommended in these high-risk populations.^18^ Additionally, female smokers have been reported to exhibit a prevalence of IAs ranging from 12%^19^ to 19%^20^.

Low ABI (≤0.9)^21, 22^ and high ABI (>1.4)^23^ serve as markers of vascular disease and can predict cardiovascular mortality beyond known cardiovascular risk factors. Low ABI correlates well with other indicators of systemic atherosclerosis, such as coronary artery calcification^24, 25^ and abdominal aortic calcification^25^, and it does not require imaging investigation/exploration, thus making it a readily available indicator. Moreover, IAs may be associated with an increased burden of atherosclerosis.^5, 6^ However, the link between cardiovascular diseases and IAs has hitherto received relatively little attention.^7, 8^

Hypertension and smoking are shared risk factors for IAs^3, 4^ and low/borderline ABI^26^. However, in our study, only low and borderline ABI both emerged as independent risk factors for IAs. Low ABI is indicative of a combination of several different risk factors,^27^ including genetics,^27^ and serves as an objective marker of vascular disease irrespective of other risk factors^21, 22^. Low ABI may be associated with systemic inflammation and endothelial dysfunction,^28^ which could increase the risk of IAs^29^. Smoking is a major shared risk factor between IAs^3^ and low ABI^28^, which partially explains our findings. However, chronic obstructive pulmonary disease, which is often caused by chronic smoking^30^, did not emerge as a risk factor for IAs in our study.

A prospective study will be required to making recommendations about IA screening based on ABI. Nonetheless, our study data show that the prevalence of IAs in low ABI and in borderline ABI groups are exceptionally high. ABI might therefore be an easy tool to screen patients who might be at a high risk for intracranial aneurysms regardless of other risk factors. Our population was too small to draw conclusions about whether there are differences between ABI groups regarding ruptured IAs. Yet despite there being no statistically significant difference, only 0.5% of patients had ruptured IAs in the normal ABI group in contrast to the low ABI and borderline ABI groups, both of which had around 2% ruptured IAs. It is intuitive to think that a population with a high prevalence of unruptured IAs may be at higher risk for ruptured IAs.

We acknowledge the limitations of our study. It is important to note that this retrospective study may introduce potential selection bias. Out of the original study population of 2757 patients, only 32% underwent cerebrovascular imaging or had ruptured IAs, and they were included in our analysis. The reasons for undergoing imaging could vary, which might lead to a possible selection bias. However, it is also worth mentioning that we had a comprehensive population, particularly for the low ABI and normal ABI groups. Despite this, the prevalence of IAs was exceptionally high in the low and borderline ABI groups, which might suggest that selection bias alone, cannot explain our findings.

Another limitation of our study is the lack of information regarding smoking status. Smoking is a well-known risk factor for both peripheral arterial disease and IAs. Unfortunately, we did not have specific information on smoking status. However, we did collect data on chronic obstructive pulmonary disease, which is often caused by heavy smoking^30^ and can serve as an indirect indicator of smoking history.

## Conclusions

The prevalence of unruptured IAs was nearly 9-fold higher in the low ABI group and approximately 6-fold higher in the borderline ABI group compared to the normal ABI group. Notably, the prevalence of unruptured IAs in the normal ABI group was similar to that reported in the general population.

## Data Availability

The data that support the findings of this study are available from the corresponding author (D.L), upon reasonable request.

## 5. Conflict of Interest

All authors declare that they have no conflicts of interest.

## 6. Sources of Funding

This study was supported by Maire Taponen foundation (Dr. Laukka), Finnish culture foundation; Varsinais-Suomen fund (grant number 85161416) and Satakunta fund (grant numbers 75212239, 75221501) (Dr Hakovirta), a government research grant awarded to Turku University Hospital (Dr. Laukka, Dr Hakovirta and Dr. Rinne) and a government research grant awarded to Satasairaala (Pori) (Dr Hakovirta).

## 7. Author Contributions

**Dan Laukka:** designed the study, acquired the data, analyzed and interpreted the data, drafted or revised the manuscript, Funding acquisition, Formal analysis, Writing – original draft. **Essi Kangas:** revised the manuscript, acquired the data. **Aino Kuusela:** revised the manuscript, acquired the data. **Jussi Hirvonen:** revised the manuscript, drafted or revised the manuscript, acquired the data, analyzed and interpreted the data. **Tiia Rissanen:** revised the manuscript, drafted or revised the manuscript, formal analysis, analyzed and interpreted the data. **Melissa Rahi:** revised the manuscript, drafted or revised the manuscript. **Juri Kivelev:** revised the manuscript, drafted or revised the manuscript. **Ville Rantasalo:** revised the manuscript, drafted or revised the manuscript. **Jaakko Rinne:** revised the manuscript, drafted or revised the manuscript, Funding acquisition. **Harri Hakovirta:** designed the study, acquired the data, analyzed and interpreted the data, drafted or revised the manuscript, Funding acquisition. All authors approved the version to be published.

## 8. Acknowledgments

We thank Auria Clinical Informatics for assisting in data collection. All authors declare that they have no conflicts of interest.

## 10. Disclosures

The authors have no personal, financial, or institutional interest in any of the drugs, materials, or devices described in this article.

## Notes

### Competing Interest Statement

The authors have declared no competing interest.

### Summary of Updates

Abstract revision.

